# Estimating duration in sex work from cross-sectional studies: a Bayesian global synthesis suggests shorter durations

**DOI:** 10.64898/2026.11.03.26367114

**Authors:** Jesse Knight, Siyi Wang, Linwei Wang, Sharmistha Mishra

**Author notes:** **Ethics Approval** Not applicable; we used only published aggregate data from 2 studies: Fazito et al. (2012) STI — Tables 1 & S2: doi.org/10.1136/sextrans-2012-050647 Baral et al. (2014) PLOS ONE — Table 1: doi.org/10.1371/journal.pone.0115465. **AI** We used no “artificial intelligence” in any part of this work. **Code** All code and selected results are available at: github.com/mishra-lab/sw-duration. **Competing Interests** None declared.

## Abstract

Durations selling sex are key determinants of HIV/STI transmission dynamics and service coverage. Such durations are usually estimated from cross-sectional surveys of people actively selling sex. However, cross-sectional surveys are more likely to include individuals selling sex for longer, and survey-reported durations will be right-censored. Prior works have not adequately addressed these two competing biases. In this paper, we derive the mathematical relationship between the distribution of durations reported in cross-sectional surveys and the distribution of true durations in the population overall. These distributions will be identical if true durations are exponentially distributed; otherwise, reported durations will typically underestimate the variance in true durations. We develop a parametric Bayesian model of this relationship, and apply it to re-analyze aggregate data from a global systematic review. Our model-estimated average true durations selling sex are significantly shorter than previously published for all regions — e.g. in Africa: 2.1 (95% CI: 1.8–2.5) versus 5.5 years; in Asia: 1.4 (1.2–1.5) versus 2.8–4.0 years. Shorter average durations then suggest that HIV incidence in sex work could be higher than previously thought, to match the same HIV prevalence data. Our approach can be applied beyond sex work, to characterize duration distributions for any transient risk/disease state.

## 1 Introduction

Epidemiological studies and healthcare programs are often interested in quantifying distributions of durations, such as durations of transient health states or exposure to risk/protective factors. Estimates of these distributions can help characterize a population’s cumulative exposure [1], and inform inputs to state-transition simulation models [2].

One prominent example is duration in sex work — selling sex for money or equivalent — reflecting a transient period of elevated risk for acquisition and transmission of HIV and other sexually transmitted infections (STI) due to intersecting structural, behavioural, and biological factors [3–5]. While entry into sex work is driven primarily by economic and social instability, exit is further mediated by aging, stable partnering, stigma, and violence [6–8]. Simulation models of HIV/STI transmission have highlighted how unmet prevention and treatment needs within sex work often drive epidemic emergence and persistence [9–11]. In such models, rates of exit from sex work are typically defined as the inverse of the mean duration in sex work — e.g. a 5-year mean duration corresponding to a 20% exit rate per year. If the sex worker population size is approximately stable over time, then exiting sex workers must be replaced by a similar number of entering sex workers — e.g. corresponding to 20% “turnover” [12].

Mean durations in sex work are thus key determinants of HIV/STI epidemiology, and key inputs to transmission models, particularly those aiming to compare the impacts of different prioritized services. For example, 10% HIV prevalence among sex workers in a context with 1-year mean duration (rapid turnover) suggests a high HIV incidence rate, whereas 10% HIV prevalence in a context with 10-year mean durations (slow turnover) suggests a lower HIV incidence rate, since prevalence has more time to accumulate [12, 13].^1^ Turnover can also moderate levels of program / service coverage among sex workers (e.g. PrEP), where rapid turnover could create a constant influx of new sex workers to reach, and potentially high loss-to-follow-up among those exiting sex work, whereas slower turnover could make it easier to attain higher levels of coverage. Finally, duration in sex work can further influence onward transmission risk, where longer durations theoretically allow more onward transmission following acquisition (in the absence of treatment).

Fazito et al. [14] conducted a global scoping review and regional syntheses of mean duration in sex work, with estimates ranging from 2.8 years in Asia to 12 years in Latin America. This review focused on cross-sectionally reported durations, and applied three methods to estimate mean duration, as data permitted: 1. mean duration among former sex workers; 2. inverse of the proportion who started sex work within the past year among active sex workers; and 3. double the mean/median duration among active sex workers, to account for right-censoring. Yet, as detailed in Table 1, each of these methods has challenges/limitations — mainly related to: difficulty sampling the target population (methods 1,2) [15], and overlooking length-biased sampling (methods 2,3) [16], as we will discuss further herein. Only some of these challenges/limitations were acknowledged in [14]. Notably, some studies also report the proportions of respondents selling sex for pre-specified duration ranges, e.g. {0–2, 2–5, 5+} years, but it’s not clear if/how such data were used in [14].

**Table 1:** Methods of estimating mean duration in sex work applied in [14].

|  | Definition | Sample | Challenges & Limitations | N<sup>†</sup> |
|---|---|---|---|---|
| 1. | mean total duration | former sex workers | • very difficult to sample this population | 0 |
| 2. | inverse of the proportion who started selling sex within the past year | active sex workers | • this proportion is rarely reported<br>• sensitive to undersampling early sex work<br>• not adjusted for length-biased sampling<sup>‡</sup> | 16 |
| 3. | double the mean/median (right-censored) duration | active sex workers | • median < mean for skewed distributions<br>• not adjusted for length-biased sampling<sup>‡</sup> | 93 |

<sup>†</sup> Number of estimates of duration selling sex synthesized by [14]; corresponding numbers were: 0, 1, 8 for duration buying sex.
<sup>‡</sup> More likely to sample individuals with longer durations [16].

Considering these challenges/limitations, we posit that estimates of mean duration in sex work reported in [14], and elsewhere using similar methods, could be substantially biased. Indeed, there appears to be uncertainty in general regarding how durations reported in cross-sectional studies relate to true overall durations in sex work. As one recent HIV modelling study noted when specifying a duration parameter’s prior distribution upper/lower bounds: *“We multiply the lower bound by 0*.*5 and upper bound by 2 because of uncertainty in how overall duration of sex work relates to current duration of sex work”* [17].

### Objectives

To address this uncertainty and challenges/limitations of existing methods to estimate mean duration in sex work, we sought to:

1. Derive the mathematical relationship between the distribution of observed durations in sex work sex from cross-sectional studies and the distribution of true durations in sex work overall.
2. Develop and apply a Bayesian model of this relationship with aggregate data from a single illustrative study [18] and a global scoping review [14], and compare model-estimated mean durations in sex work to previously published estimates.

The remainder of our paper is structured as follows: In § 2, we clarify some key concepts and assumptions for the work. In § 3, we derive the mathematical relationship for Objective 1. In § 4, we develop and apply the Bayesian model for Objective 2. In § 5, we conclude with discussion of implications and future work.

## 2 Preliminaries

### Assumptions

For simplicity, we first assume that periods of sex work have clear start/end points, and that formal sex work can be clearly distinguished from transactional sex. We also assume that surveys predominantly reach those actively engaged in sex work, despite some survey eligibility criteria including individuals who sold sex anytime in the past year, etc. [3]. We further assume that survey participants can accurately report how long they have sold sex — i.e. perfect recall, no social desirability bias. Finally, we assume that all populations are at equilibrium, e.g. rates of entering/exiting sex work are stable over time.

### Who exactly are we talking about?

Before continuing, it is helpful to distinguish two populations: individuals who will ever sell sex in their lifetime, and individuals who are actively selling sex. The former “source population” is the target sample for method 1 in [14], and tends to include more individuals who sell sex for shorter durations; the latter “active population” (a subset of the source population) is the target sample for methods 2 and 3 in [14], and tends to include more individuals who sell sex for longer durations. Figure 1 illustrates periods in sex work for a hypothetical source population at equilibrium, distinguishing the active population (green) from the inactive population (gray) at the time of a hypothetical survey (yellow).

**Figure 1:**
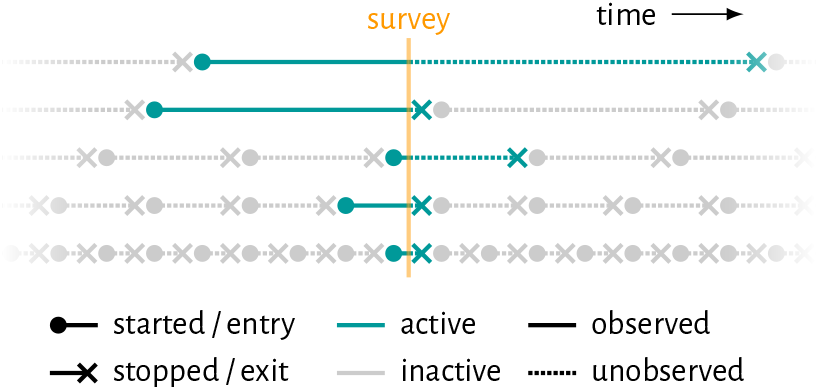
Diagram of periods selling sex in an open population at equilibrium. Note: consecutive periods in each row denote multiple individuals, not multiple periods of sex work by the same individual.

Since “sex work” strata in compartmental models represent individuals actively selling sex, we might assume that rates of exit from these strata should be derived from durations among the active population. However, such rates should actually be derived from durations among the source population, because exit rates apply to all individuals *entering* sex work, which reflects the source population. In an analogous example: the duration of an infection should be characterized by a cohort of individuals with *incident* infections, not a cross-section of individuals with *prevalent* infections.

A final population worth distinguishing is the “sampled population” in a given cross-sectional survey, which we assume intends to be representative of the active population, but might undersample some individuals, e.g. those in the early years of sex work, due to delays in self-identification as a sex worker [19, 20].

## 3 Mathematical relationships

We now turn to deriving the mathematical relationship between: the distribution of true/total durations in sex work among the source population: *x ∼ f* (*x*), and the distribution of observed/censored durations in sex work among participants in a cross-sectional survey: *z ∼ f* (*z*|*s*), given survey participation status *s*. For reference throughout this section, Table A.1 summarizes our notation, while Figure 2 illustrates example distributions for *f* (*x*), *f* (*z*|*s*), and intermediates thereof.

**Figure 2:**
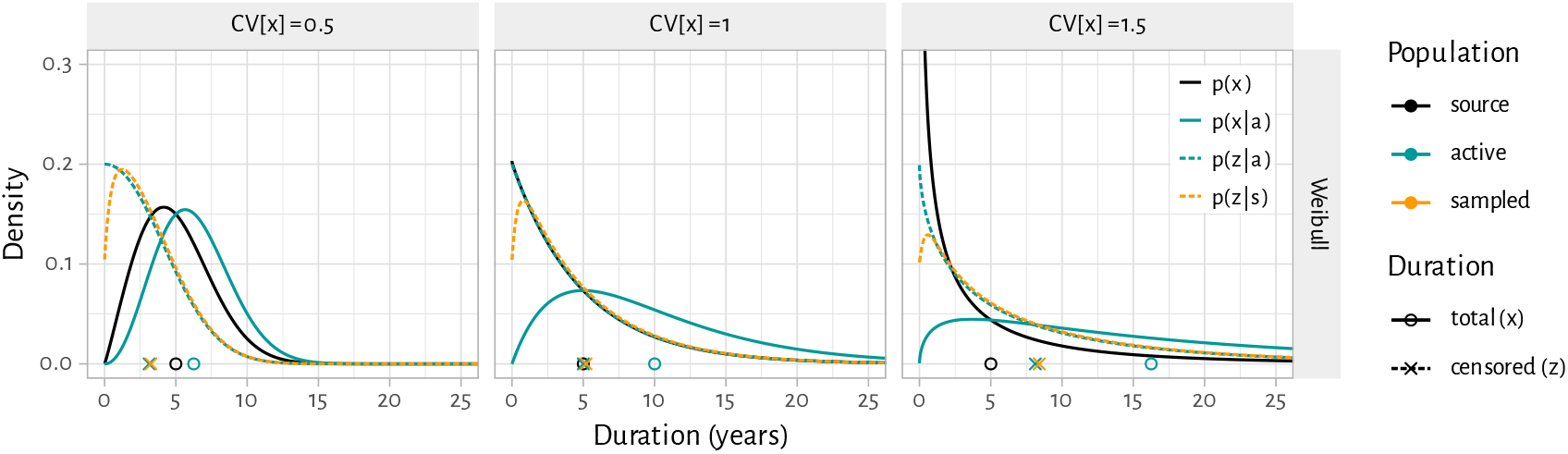
Illustrative distributions of true/total and observed/censored durations among the source, active, and sampled populations for 3 coefficients of variation ℂV[*x*] (columns) Assumed: mean E[*x*] = 5 and *x ∼* Weibull (Figure A.1 illustrates other distribution families); probability of being sampled per Eq. (10).

### Length-biased sampling

The probability of an individual being active at any given time is proportional to their total duration *x*; a phenomenon well-described (e.g. in cancer epidemiology and renewal theory) as “length-biased sampling” or the “renewal paradox” [16, 21]. Thus, the distribution of total durations among the active population is [21, 22]:

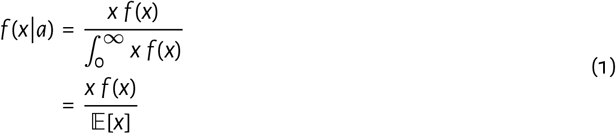

where E[*x*] is the mean total duration among the source population. These total durations among the active population *x*|*a* are typically unobserved, except in cohort studies which follow participants until they exit sex work. From Eq. (1) we can show that the mean total duration among the active population E[*x*|*a*] is always larger than the mean among the source population E[*x*] (Figure A.1, green vs black circles); specifically [23]:

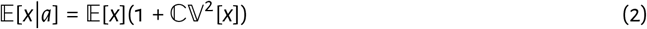

This is because greater variability in durations among the source population, as measured by the coefficient of variation: ℂ V[*x*], allows those with longer durations to increasingly dominate the active population. For reference, we expect that ℂV[*x*] *∈* (1, 2) in this context, for three reasons. First, heavy-tailed distributions having ℂV[*x*] > 1 can arise via selection effects in a population with heterogeneous exit rates [24]. Second, most parametric distributions with ℂV [*x*] < 1 have *f* (*x →* 0) *→* 0 (Figure A.1), which implies that few people exit sex work shortly after starting, or that most people stay in sex work for a “minimum” period, which seems unrealistic. Finally, aging naturally constrains maximum durations, which in turn constrains duration variance, such that usually ℂV [*x*] < 2.

Returning to Eq. (1), we also note that the probability of observing active individuals with short total durations is inherently small (Figure 2, green solid line). Relatedly, the *inactive* population (denoted *ā*), includes many future or former sex workers with short total durations (Figure 1, gray), and thus has smaller mean total duration than the source or active populations:E[*x*|*ā*] < E[*x*] < E[*x*|*a*].

### Censoring

Next, we consider a hypothetical survey among the active population. If timing of the survey is effectively random, then observed durations at survey time (*z*) will be right-censored and uniformly distributed between 0 and the total duration (*x*):

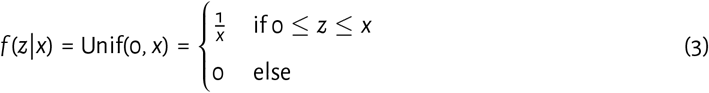

Since the mean of Unif(0, *x*) is 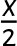, the mean censored duration will be half the mean total duration among the active population (Figure 2, green crosses vs circles):

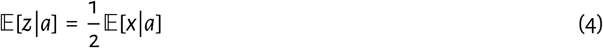

This idea appears to be well-known, and likely motivates doubling the mean E[*z*|*a*] to estimate E[*x*] in method 3 of [14]. However, method 3 does not account for length-biased sampling, and thus actually estimates the mean total duration among the active population E[*x*|*a*], which we know to be greater than among the source population E [*x*] per Eq. (2). ^2^

### Complete distribution

To consider both length-biased sampling and censoring simultaneously, we can combine Eqs. (2) and (4) to relate the mean total duration among the source population E[*x*] to the mean censored duration among the active population IE[*z*|*s*]:

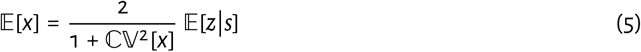

However, this is not immediately useful unless ℂV[*x*] is known. Alternatively, by combining Eqs. (1) and (3), we can relate the complete distributions *f* (*x*) and *f* (*z*|*a*) — Eq. (A.1) gives a complete derivation [22]:

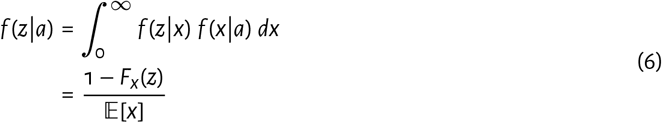

As it turns out, this is simply the “survival function” or “tail distribution” of total durations *x* (one minus the cumulative distribution function *F*_*x*_), normalized to have unit area under the curve via E[*x*] as in Eq. (1). Since this cumulative distribution is initially *F*_*x*_ (*z* = 0) = 0 by definition, we can estimate E[*x*] as the inverse of the proportion of active sex workers who “just started” (Figure 2, y-intercept of green dashed lines):^3^

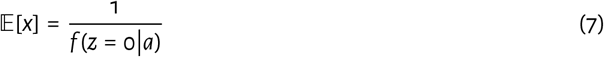

We can then recover *f* (*x*) by differentiating *f* (*z*|*a*) and multiplying by E[*x*]:

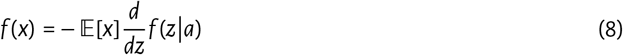

Eq. (8) is only useful if the complete distribution *f* (*z*|*a*) is known, or can be approximated e.g. via histogram. If only summary statistics for *f* (*z*|*a*) are available, we can still leverage Eq. (6) to infer *f* (*x*), as we show in § 4.

### Exponential case

A special case arises if durations are exponentially distributed: *f* (*x*) = *λe*^−*λx*^, which is often implicitly assumed in compartmental simulation models [2]. Since E[*x*] = *λ* ^−1^ and *F*_*x*_ = 1 – *e*^−*λx*^, we find that: *f* (*z*|*a*) = *f* (*x*). That is, the distributions of *total* durations among the *source* population and *censored* durations among the *active* population will be exactly equal. Under these conditions, the mean reported duration in a typical cross-sectional survey E[*z*|*a*] will directly estimate the target mean duration E[*x*]. This equality is also indicated by Eq. (5), since ℂV[*x*] = 1 for the exponential distribution. However, as noted above, often ℂV[*x*] > 1, in which case E[*z*|*a*] would overestimate E[*x*].

### Further sampling bias

Until now we have assumed that surveys reach a representative sample of the active population. If a given survey further undersamples individuals in the early years of sex work — i.e. beyond effects of length-biased sampling, due to delays in self-identification as a sex worker [20] — this probability of being sampled will be a function of current observed/censored duration (*z*) not true/total duration (*x*), so we denote it *f* (*s*|*z*). Many parameterizations of *f* (*s*|*z*) are possible; e.g. Eq. (10) below in § 4. To incorporate this further sampling bias, we update the distribution of observed durations among survey participants from Eq. (6) as:

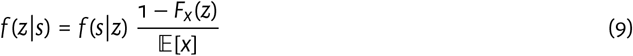

In this context, *f* (*x*) cannot be directly determined via Eq. (8), but it can be inferred as we show in § 4.

### Revisiting prior methods to estimate E[*x*]

We conclude this section by revisiting the three methods applied by Fazito et al. [14] to estimate the “true” mean duration in sex work E[*x*] (Table 1). Figure A.2 plots: the typically unobserved mean and median of total durations (method 1),^4^ the reciprocal proportion of censored durations less than 1 year (method 2), and *double* the means and medians of censored durations (method 3), across a range of ℂV[*x*] and distribution families for *f* (*x*) with true E[*x*] = 5. We see that at low ℂV[*x*], nearly all methods converge on the true E[*x*]. However, as ℂV[*x*] increases, length-biased sampling causes stronger selection towards longer total durations among the active population, raising the corresponding estimates of E[*x*] according to Eq. (2), e.g. twice as large at ℂV [*x*] = 1. Median censored durations exhibit a similar trend, but lag below means (true for all right-skewed distributions) to a degree which depends on the family, making such medians less useful estimators of E[*x*] if the family is unknown. For all method 3 estimates, undersampling of early sex work (here assuming *ρs* = 0.5, *τ*_95_ = 1 year) predictably biases estimates slightly higher; this bias is also larger for smaller E[*x*] (not shown). Finally, method 2 tends to be most consistent, only slightly overestimating E[*x*] as ℂV[*x*] increases. However, this method is especially sensitive to undersampling of early sex work, which biases estimates upwards.

## 4 Application

We implemented the mathematical relationship Eq. (9) in a Bayesian framework, allowing us to infer the distribution of total durations among the source population *f* (*x*) from individual-level and/or aggregate data on the distribution of censored durations among the sampled population *f* (*z*|*s*). We focused on the types of aggregate estimates reviewed by Fazito et al. [14]: duration means 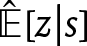, and proportions of durations within a given interval [*a, b*], denoted 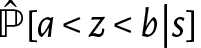 — e.g. if 33% of durations were between 2 and 5 years, then 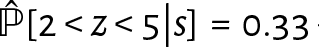— which can also encompass estimated medians, quartiles, etc. with *a* = 0.

### 4.1 Bayesian Model

The model assumes a parametric distribution for total durations among the source population *f* (*x*). Specifically, we considered 5 candidate distribution families: Exponential, Gamma, Weibull, Scaled Beta, and Scaled Kumaraswamy, all truncated or scaled to 50 years maximum duration (Table A.2). For convenience, we denote the parameters of all families as: *α, β* (for Exponential: *β* only), so the interpretation of and priors for each parameter depend on the family.

We modelled the probability of being sampled for survey participation among the active population as increasing exponentially from *ρs ∈* [0.5, 1] initially to 95% by year *τ*_95_ *∈* [0, 3], and 100% asymptotically (Figure A.3):^5^

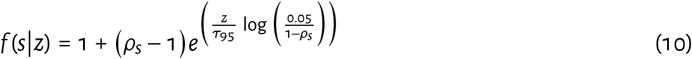

We then defined the distribution of censored durations among the sampled population *f* (*z*|*s*; *α, β, ρs, τ*_95_) via Eq. (9). Finally, Eqs. (A.2) and (A.3) define the likelihood functions for estimates 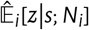 and 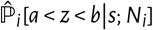 from empirical data with sample size *Ni*. Similar functions could be specified for individual-level data.

#### Implementation

We implemented the model using Stan [25], with default random parameter initialization and sampling algorithm [26]. For a given set of empirical data — i.e. aggregate estimate(s) of 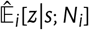 and/or 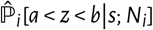 — we fit the model separately under each candidate family, and then selected the best-fitting family (highest total likelihood). We retained 1000 consecutive samples after 500 warm-up iterations from 7 chains, confirming convergence visually through parameter trace plots (e.g. Figure A.9).^6^ This approach yielded 7000 posterior parameter samples, from which we computed the distribution of total durations among the source population *f* (*x*; *α, β*), and characteristics thereof, e.g. the mean E[*x*; *α, β*], each with uncertainty.

### 4.2 Aggregate data from a single survey

We first demonstrated the model on aggregate data from a 2011 cross-sectional survey of 328 female sex workers in Eswatini [18]. The authors reported crude and respondent-driven sampling (RDS)-adjusted mean (95% CI) proportions of respondents who had been in sex work for 0–3, 3–6, 6–11, and 11+ years (Table 2) — i.e. estimates of 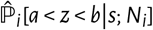. We converted these 95% CI to effective sample sizes per stratum (*Ni*), for compatibility with the likelihood function Eq. (A.3), and fit the model as described above.

**Table 2:** RDS-adjusted proportions of durations in sex work reported by a 2011 sample of female sex workers in Eswatini (N = 313) [18].

| Duration<sup>†</sup> | Proportion (%) |  |  | *N<sub>i</sub>* |
|---|---|---|---|---|
| (years) | Crude | RDS | (95% CI) | Eff. <sup>‡</sup> |
| 0–3 | 28.8 | 38.3 | (27.5, 49.1) | 77 |
| 3–6 | 31.3 | 32.1 | (23.6, 40.7) | 113 |
| 6–11 | 26.2 | 20.2 | (13.2, 27.1) | 129 |
| 11+ | 13.7 | 9.4 | (4.4, 14.4) | 135 |

<sup>†</sup> Durations were calculated as the difference between current age and age first reported selling sex; duration strata in [18] were denoted: "under 2, 3–5, 6-10, 11 or more", which we corrected here to avoid implied gaps between strata. <sup>‡</sup> Effective sample size per strata calculated to match RDS-adjusted 95% CI.

The best-fitting family was Gamma. We found that the model could reproduce the empirical proportion data well (Figure 3.i), but may have suffered from unidentifiability using these data alone — i.e. many combinations of parameters could reproduce the data equally well. For example, posterior uncertainty was low around the observed duration distribution *f* (*z*|*s*), but high around the inferred “true” duration distribution *f* (*x*) (Figure 3.ii, colour vs gray). Similarly, posterior 95% CI were relatively narrow for E[*z*|*s*]: 4.37–5.83 and ℂV[*z*|*s*]: 0.72–1.15, but wide for E[*x*]: 2.30–8.53 and ℂV[*x*]: 0.43–1.79 (Figure A.8). Posteriors for E[*z*|*s*] and ℂV[*z*|*s*] also remained similar across families, whereas posteriors for E[*x*] and ℂV[*x*] varied even further across families (Figure A.8).

**Figure 3:**
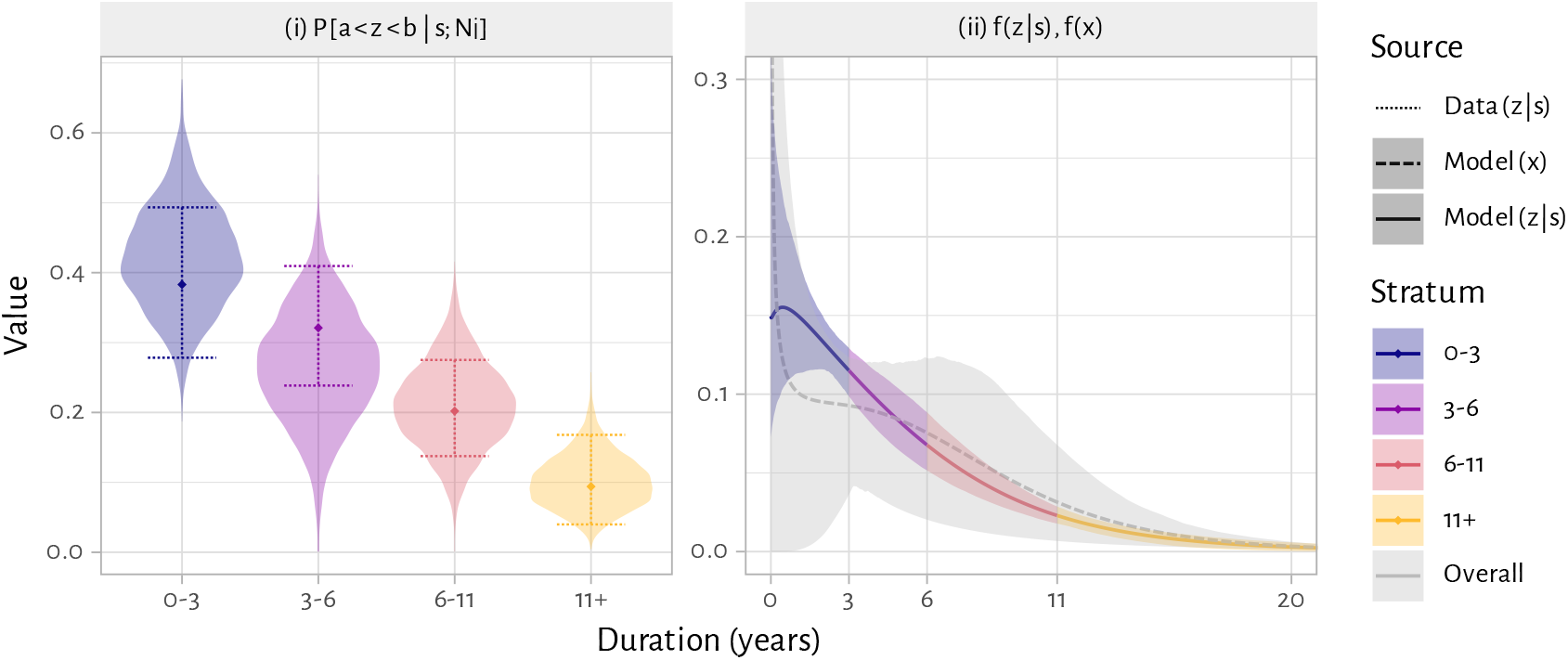
Posterior estimates from fitting the Bayesian model to RDS-adjusted aggregate censored duration data from [18]: (i) proportions of sampled respondents with durations in pre-specified intervals ℙ[*a* < *z* < *b*|*s*; *N*_*i*_]; (ii) inferred distributions of: censored durations among sampled population *f* (*z*|*s*) (colour), and total durations among source population *f* (*x*) (gray) Shaded regions denote: (i) posterior distribution of modelled proportions given effective sample size *N*_*i*_; (ii) posterior 95% CI of modelled duration distribution density. Points and dashed line ranges denote RDS-adjusted means and 95% CI for proportion data in (i).

This identifiability issue likely arose because the data aggregated the first 0–3 years into one stratum. This especially limited our ability to infer parameters of the early sex work sampling model: *ρ*_*s*_, *τ*_95_, whose posterior distributions were effectively indistinguishable from their priors (Figure A.8). Nevertheless, we interpret our inference regarding the “true” duration distribution *f* (*x*) as conservatively realistic regarding remaining uncertainty, based on these empirical data alone.

### 4.3 Aggregate data from a global review

Next, we applied our model to generate regionally pooled estimates of mean total duration in sex work using the same aggregate data and region definitions as Fazito et al. [14], and compared our pooled estimates to theirs.^7^ We manually extracted aggregate data for female sex workers from Table S2 in [14] on duration means 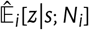 and interval proportions 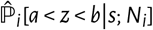, including survey region, sample size *N*_*i*_, and other meta-data (see § A.2.2 for details). We then fit our model separately for each region with data, retaining the best-fitting family per region. Figures A.10–A.14 illustrate the posterior distributions of model parameters, summary statistics, cumulative/density duration distributions, and means versus the data, stratified by region (Figure) and family (colour).

Figure 4 then illustrates model-estimated mean total durations IE[*x*] (best-fitting family per region) versus those in Table 1 of [14]. Our estimates are significantly shorter than those of [14] for all regions — e.g. we estimated: 2.1 (1.8–2.5) versus 5.5 years for Africa (approximately 2.5x shorter), 1.4 (1.2–1.5) versus 2.9 years for Asia (approximately 2x shorter), and 2.9 (2.0–3.9) versus 11.2–12.0 years for Latin America (approximately 4x shorter). These differences were expected when considering that method 3 can overestimate E[*x*] by failing to account for length-biased sampling, while method 2 can overestimate E[*x*] if early sex work is undersampled. Model-estimated distributions of total durations among the source population *f* (*x*) were also consistently heavy-tailed, with ℂV[*x*] > 1.5, suggesting substantial population heterogeneity [24]. Relatedly, the model estimated that over half of all source population women remain in sex work for less than 1 year in all regions except North America, which had only 3 studies (Figures A.10–A.14).

**Figure 4:**
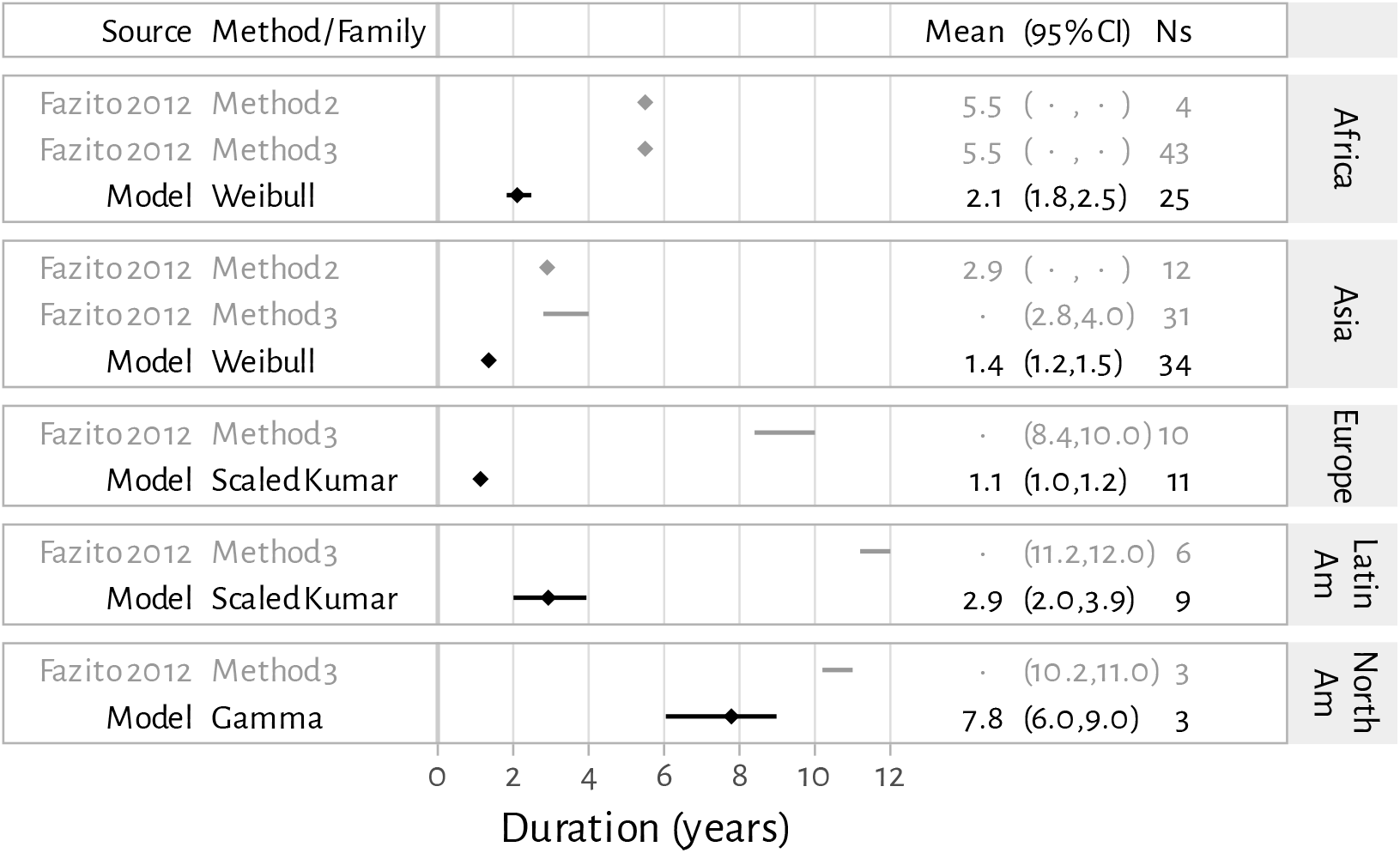
Forest plot of Bayesian model-estimated mean total duration in sex work IE[*x*] (best-fitting family per region) versus regional synthesis estimates in [14]. Synthesis methodology in [14] was unclear, and regional estimates were variably reported as points or ranges; see Table 1 for method definitions. Ns: number of unique surveys contributing to the estimate.

## 5 Discussion

In this paper, we have clarified several concepts and challenges relevant to characterizing the distribution of durations in sex work from cross-sectional data. First, we distinguished between “source” and “active” populations, with the active population being a length-biased subset of the source population, where the probability of being active is proportional to total duration in sex work. Second, we reiterated the familiar implications of randomly censored durations, which halves the mean duration. We combined the above to derive the exact mathematical relationship between the distribution of censored durations among the active population and the distribution of total durations among the source population, which we showed to be equal if durations are Exponentially distributed. We then developed a parametric Bayesian model of this relationship to infer the distribution of total durations among the source population from aggregate data on censored durations among the active population. Applying this model to data from a previous global systematic review [14], we inferred significantly shorter mean durations across multiple regions.

Our estimates of mean duration in sex work were significantly less than those reported by Fazito et al. [14]. This was expected, as we showed how both methods 2 and 3 can overestimate mean duration, especially if surveys undersample individuals in early sex work. Shorter durations means that that incidence of HIV and other STI in sex work could be higher than previously estimated via transmission models, since there is less time for infections to accumulate to match observed HIV/STI prevalence in sex work [12, 13]. However, existing model-based estimates of HIV incidence among female sex workers in Sub-Saharan Africa already exceed empirical estimates (mainly from cohorts and trials) [27, 28].

Shorter durations, and thus more rapid sex work turnover, could also suggest that a greater proportion of women in the wider population might have previously sold sex. Indeed, quasi-anonymous polling booth surveys have indicated substantial under-reporting lifetime sex work engagement for both women and men in face-to-face interviews [29, 30]. A greater proportion of prevalent infections in the population overall might then be attributable to brief, unreported periods of sex work, additionally considering unique vulnerabilities of early sex work [20]. We reiterate that such brief periods in sex work are especially difficult to observe in cross-sectional studies due to length-biased sampling [21]. Taken together, these findings underscore the need to rapidly identify and reach women during early sex work with relevant programs and services, e.g. through place-based strategies, to efficiently reduce population-level HIV/STI burden [20, 31–33].

We also inferred that distributions of total durations in sex work overall are likely “heavy-tailed”, such that rates of exit decline with longer duration in sex work. This declining exit rate could reflect selection effects acting on fixed between-person heterogeneity in a propensity to remain in sex work [24], and/or within-person changes that make it easier to remain in sex work. including self-identification as a sex worker, and changing social ties with family, friends, and clients, [6–8, 31, 32]. While individual-based simulation models could specify such heterogeneity or declining exit rates directly, compartmental simulation models would need two or more sex work states to reproduce this heavy-tail — e.g. separate short and long duration sex work trajectories, and/or separate early and late sex work periods. Indeed, such states would also be useful to capture unique risks associated with early sex work [31, 32].

We focused on cross-sectional studies, which have been the cornerstone of sexual health epidemiology globally [34, 35], since they are generally simpler and less expensive than cohort studies [14]. However, a cohort design does not necessarily simplify estimation of “true” sex work durations, as sampling can still be length-biased (depending on how participants are recruited) and significant loss-to-follow-up can occur [15]. While the proposed Bayesian model can leverage multiple kinds of aggregate data on durations, future cross-sectional studies could most helpfully report complete duration histograms, where possible, and the following key summary statistics: minimum, maximum, mean, median, and interquartile range of durations, as well as the proportion of active individuals in sex work 1 year or less.

### Limitations

Our work has several limitations. First, we did not model any gaps (temporary breaks) in sex work. Many estimates of sex work duration are derived from the difference between respondents’ current age and the age they report first selling sex, which implicitly ignores such gaps. Respondents may also report “duration in sex work” in this way when asked directly, especially since unsuccessful attempts to exit sex work can be stigmatized [8]. Second, we did not consider any recall/reporting biases, as entry into sex work is likely a memorable life-course event [6–8, 36, 37], and self-identified active sex workers do not, to our knowledge, have obvious motivations for under-reporting durations. We additionally assumed that sex work turnover is stable over time, despite empirical evidence of changing sex work dynamics in several contexts [13, 38]. Finally, our re-analysis of duration data from [14] did not include more recent or individual-level data, since our motivation was mainly to compare results using the Bayesian model versus prior estimation methods.

### Future work

Our Bayesian model could be extended in several ways. First, it might be more useful to model sex work durations via semi/parametric hazard rates of exiting sex work, rather than parametric duration distributions. A hazard rate approach could allow and incorporation of more intuitive parameter priors (e.g. 0.5–50% exit rate per month), covariates (e.g. under proportional hazards assumptions) [39], and frailties (e.g. to capture unmodelled heterogeneity) [24]. Relatedly, joint modelling of duration and age and/or calendar time could help characterize relevant trends noted above e.g. for HIV/STI transmission modelling [13, 38]. Such extensions might be supported by related work estimating durations (also called sojourn times) in chronic disease states for modelling and analysis of screening programs [23].

### Conclusion

The distribution of durations in sex work is a key determinant of HIV/STI dynamics and an essential input to HIV/STI transmission models. Such durations are commonly reported in cross-sectional studies, where they are subject to two competing biases: length-biased sampling (increases the mean) and censoring (decreases the mean). The former bias, omitted from prior work, is likely stronger, such that the mean duration reported in cross-sectional studies actually overestimates rather than underestimates the “true” mean duration. Methods herein also support inference of the complete distribution of “true” durations, given a complete distribution of reported durations. Notably, these methods can be applied to characterize the distribution of durations beyond sex work, such as for transient health states, or periods of exposure to risk/protective factors.

## Data Availability

We used only published aggregate data. All code and selected results are available on GitHub

https://github.com/mishra-lab/sw-duration

## Acknowledgements

We thank: Oli Stevens, Rebecca Anderson (Imperial College London), Nancy Tahmo, Chidumebi Idemili (Unity Health Toronto) for helpful discussions regarding this work; as well as two anonymous reviewers for feedback on an early precursor to this work.

## Footnotes

1 Assuming HIV prevalence among new sex workers is low.

2 Fazito et al. [14] partially acknowledge this issue by opting to use the median over the mean of *z*|*a* where possible, since the median is less sensitive to outliers. However, as shown in Figure A.2, doubling the median M[*z*|*a*] still overestimates E[*x*], just to a lesser degree than doubling the mean E[*z*|*a*], since the median will always be less than the mean for right-skewed distributions.

3 Eq. (7) is essentially a more precise definition of method 2 in [14].

4 We are being generous to method 1 here by assuming former sex workers are representative of the *source* population, whereas only the *inactive* population is sampled in practice.

5 Thus, if either *ρ*_*s*_ = 1 or *τ*_95_ = 0, then sampling will be modelled as representative of the active population: *f* (*s*|*z*) = 1 and so *f* (*z*|*s*) = *f* (*z*|*a*).

6 We manually removed 3 chains that did not converge.

7 We acknowledge that region definitions in [14] are now outdated, e.g. combining all of Africa.

