## Appendix for "Estimating duration in sex work from cross-sectional studies: a Bayesian global synthesis suggests shorter durations"

**Date** November 2, 2026

### Appendix A

### Supplementary Content

Table A.1: Notation reference

| Symbol | Definition |
|---|---|
| $x$ | true/total duration |
| $z$ | observed/censored duration |
| $\cdot$ | duration among source population (unconditional) |
| $\cdot \mid a$ | duration among active population |
| $\cdot \mid k$ | duration among sampled population |
| $f(\cdot)$ | density distribution |
| $F(\cdot)$ | cumulative distribution |
| $K$ | a normalizing constant |
| $\mathbb{E}[\cdot]$ | mean |
| $\mathbb{M}[\cdot]$ | median |
| $\mathbb{CV}[\cdot]$ | coefficient of variation (standard deviation / mean) |
| $\mathbb{P}[\cdot]$ | probability |
| $\alpha, \beta$ | distribution parameters (depends on family: see Table A.2) |
| $\rho_s$ | probability of being sampled immediately after entry |
| $\tau_{95}$ | duration by which probability of being sampled is 95% |
| $N_i$ | sample size for empirical estimate $i$ |

#### A.1 Mathematical relationships

**Derivation of $f(z|a)$** Eq. (A.1) gives the complete derivation of Eq. (6) — i.e. the complete distribution of censored durations among the active population $f(z|a)$ from the distribution of total durations among the source population $f(x)$ [1]:

$$\begin{aligned} f(z|a) &= \int_0^\infty f(z|x)\, f(x|a)\, dx \\ &= \int_z^\infty \frac{1}{x}\, f(x|a)\, dx \\ &= \int_z^\infty \frac{x}{x} \frac{f(x)}{\mathbb{E}[x]}\, dx \\ &= \frac{1}{\mathbb{E}[x]} \left(1 - F_x(z)\right) \end{aligned} \tag{A.1}$$

Note that the integral lower bound changes from 0 to $z$ because $f(z|x) = \frac{1}{x}$ only when $x < z$ and $f(z|x) = 0$ otherwise, per Eq. (3).

**Distribution families** We considered 5 distribution families for the true/total durations among the source population $f(x)$ in Objectives 1 & 2 — Table A.2 defines these families using our unified notation, and specifies parameter priors used for the Bayesian model.

Table A.2: Distribution families considered, using unified parameter notation $(\alpha, \beta)$

| Family | Definition $f(x)$ | $\alpha$ log-prior * | $\beta$ log-prior * |
|---|---|---|---|
| Exponential † | $\frac{1}{\alpha}\, e^{-\frac{x}{\alpha}}$ | – | [–1, 3] |
| Gamma † | $\frac{\beta^{\alpha}}{\Gamma(\alpha)}\, x^{\alpha-1} e^{-\beta x}$ | [–2, 2] | [–3, 3] |
| Weibull † | $\frac{\alpha}{\beta}\left(\frac{x}{\beta}\right)^{\alpha-1} e^{-(x/\beta)^{\alpha}}$ | [–1, 2] | [–1, 2] |
| Scaled Beta ‡ | $\frac{x^{\alpha-1}(1-x^{\alpha})^{\beta-1}}{B(\alpha,\beta)}$ | [–3, 3] | [0, 3] |
| Scaled Kumaraswamy ‡ | $\alpha\,\beta\, x^{\alpha-1}(1-x^{\alpha})^{\beta-1}$ | [–3, 3] | [0, 3] |

* Priors for $\alpha, \beta$ in the applied model were defined as uniform on the log-scale; e.g. [–1, 3] corresponds to $[e^{-1}, e^{3}] \approx [0.37, 20]$.
† Distributions spanning $[0, \infty)$ were truncated to [0, 50] years. ‡ Distributions spanning [0, 1] were scaled to span [0, 50] years.

**Illustrative analytic results** Figure A.1 repeats Figure 2 for 5 distribution families considered (Table A.2) — i.e. showing distributions of total ($x$) and censored ($z$) durations among the source, active, and sampled populations for each family, all having $\mathbb{E}[x] = 5$ and $\mathbb{CV}[x] = \{0.5, 1, 1.5\}$. We omit the Exponential family as it is a special case of both Gamma and Weibull with $\mathbb{CV}[x] = 1$. Figure A.2 further illustrates performance of different $\mathbb{E}[x]$ estimators considered in [2] across a range of $\mathbb{CV}[x]$ and the same 5 distribution families. Finally, Figure A.3 illustrates the shape and parameters of the assumed sampling model Eq. (10).

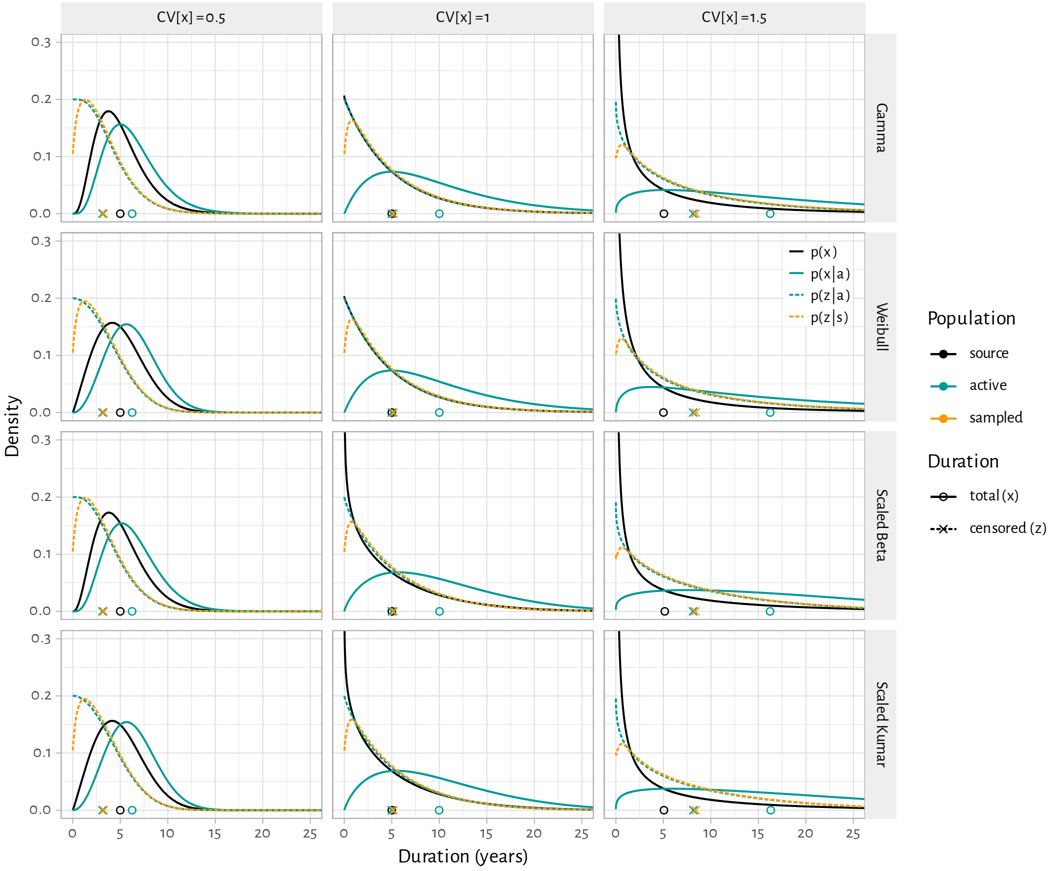


Figure A.1: Illustrative distributions of total and censored durations among the source, active, and sampled populations for different $x$ distribution families (columns) and 3 coefficients of variation $\mathbb{CV}[x]$ (rows)

Note: true $\mathbb{E}[x] = 5$; probability of being sampled modelled per Eq. (10) with $\rho_s = 0.5$, $\tau_{95} = 1$.

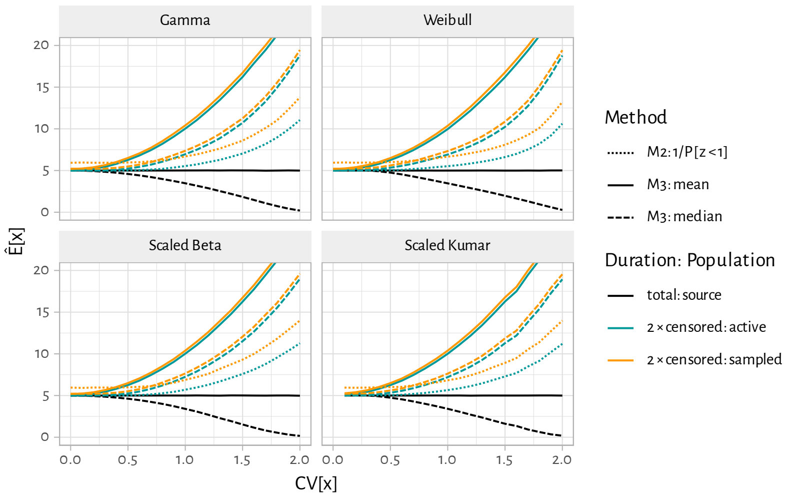


Figure A.2: Performance of $\mathbb{E}[x]$ estimators considered in [2] across a range of $\mathbb{CV}[x]$ and distribution families

Note: true $\mathbb{E}[x] = 5$; probability of being sampled modelled per Eq. (10) with $\rho_s = 0.5$, $\tau_{95} = 1$; means and medians for censored data ($z$) are doubled per [2].

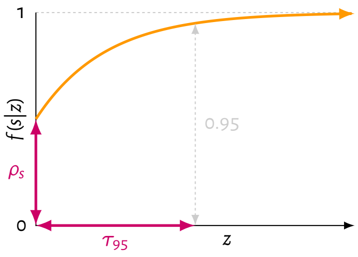


Figure A.3: Diagram of $f(s|z)$ per Eq. (10): modelled probability of being sampled versus current duration $z$

Parameters: $\rho_s$: initial probability of being sampled immediately after entering sex work; $\tau_{95}$: duration by which probability of being sampled reaches 95%.

#### A.2 Application

**Likelihoods functions** Under the model for $f(z|s)$, we define the likelihoods for empirical estimates of the mean $\hat{\mathbb{E}}_i[z|s]$ and proportions $\hat{\mathbb{P}}_i[a < z < b|s]$ from a given survey with sample size $N_i$ as, respectively: the Gamma-equivalent of Normal sampling error for the mean (since $\hat{\mathbb{E}} > 0$), and the Beta-approximation of the Binomial distribution (for differentiability):

$$\hat{\mathbb{E}}_i[z|s; N_i] \sim \text{Gamma}(\alpha = N_i \,/\, \mathbb{CV}_i^2[z|s],\;\; \beta = N_i \,/\, \mathbb{CV}_i^2[z|s] \,/\, \mathbb{E}[z|s]) \tag{A.2}$$

$$\hat{\mathbb{P}}_i[a < z < b|s; N_i] \sim \text{Beta}(\alpha = N_i \, \mathbb{P}_i[a < z < b|s],\;\; \beta = N_i \, (1 - \mathbb{P}_i[a < z < b|s])) \tag{A.3}$$

To validate these likelihood functions, we simulated 10,000 stochastic samples of $z$ with sample sizes $N_i = \{30, 100, 300, 1000\}$ given known $f(z|s)$ under $\mathbb{E}[x] = 5$, $\mathbb{CV}[x] = \{0.5, 1, 1.5\}$, and all 6 distribution families. We then compared the distribution of stochastic estimates of $\hat{\mathbb{E}}[z|s; N_i]$ and $\hat{\mathbb{P}}[a < z < b|s; N_i]$ from each sample with the analytic likelihood functions: Eqs. (A.2) and (A.3). For proportions, we considered $\mathbb{P}[0 < z < 1]$, $\mathbb{P}[1 < z < 5]$, and $\mathbb{P}[5 < z < 9]$, arbitrarily.

Figures A.4–A.7 illustrate these comparisons, where we observe good agreement between the stochastic and analytic distributions across all conditions. Note that stochastic distributions for $\hat{\mathbb{P}}[a < z < b|s; N_i]$ are discrete (more visible at low $N_i$) because there are a finite number simulated respondents whose durations can fall in the specified interval $[a, b]$.

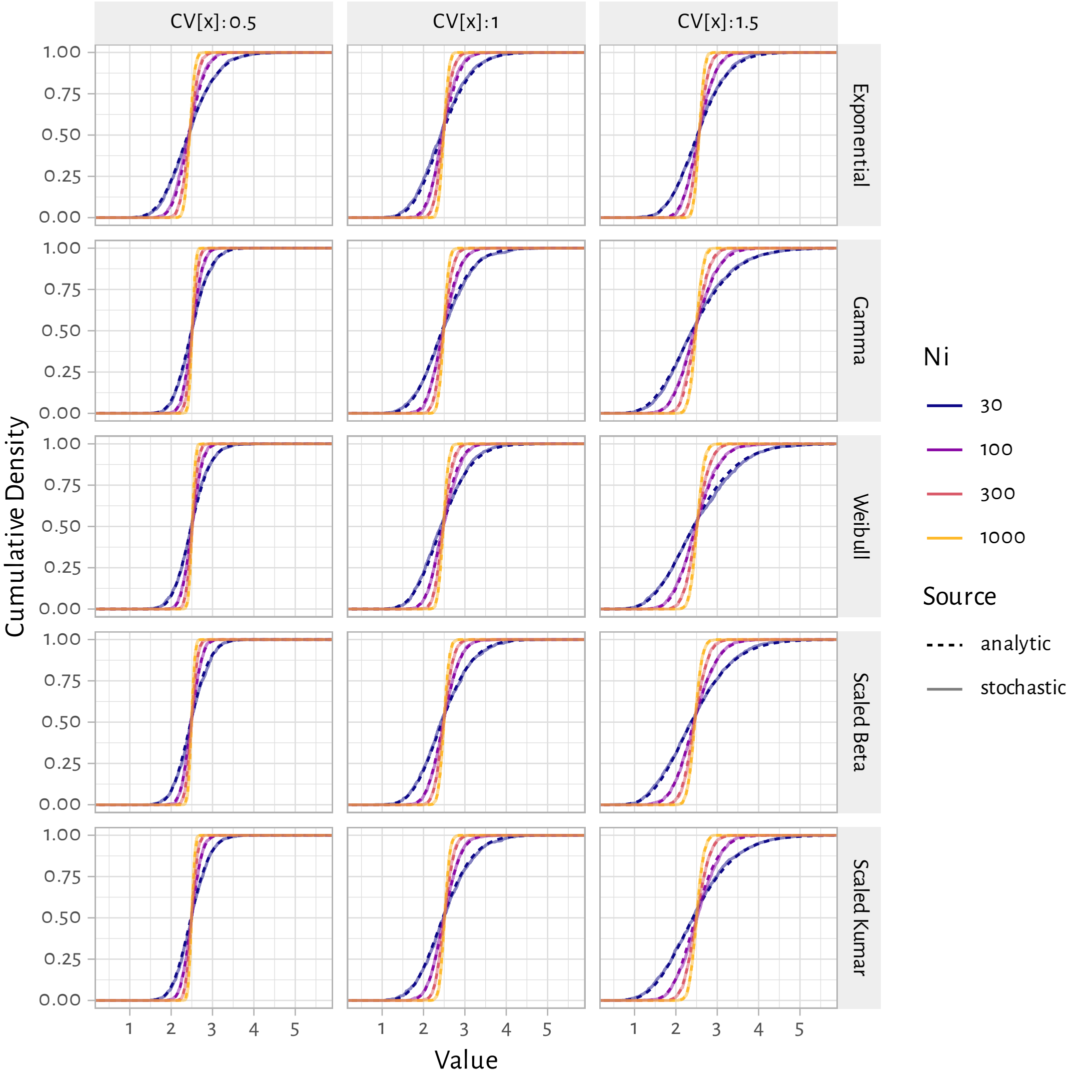


Figure A.4: Comparison of analytic likelihood functions and distribution of 10,000 stochastic estimates of $\mathbb{E}[\boldsymbol{z}|\boldsymbol{s}]$, for different simulated survey sizes $N_i$ and latent distribution families for $f(x)$

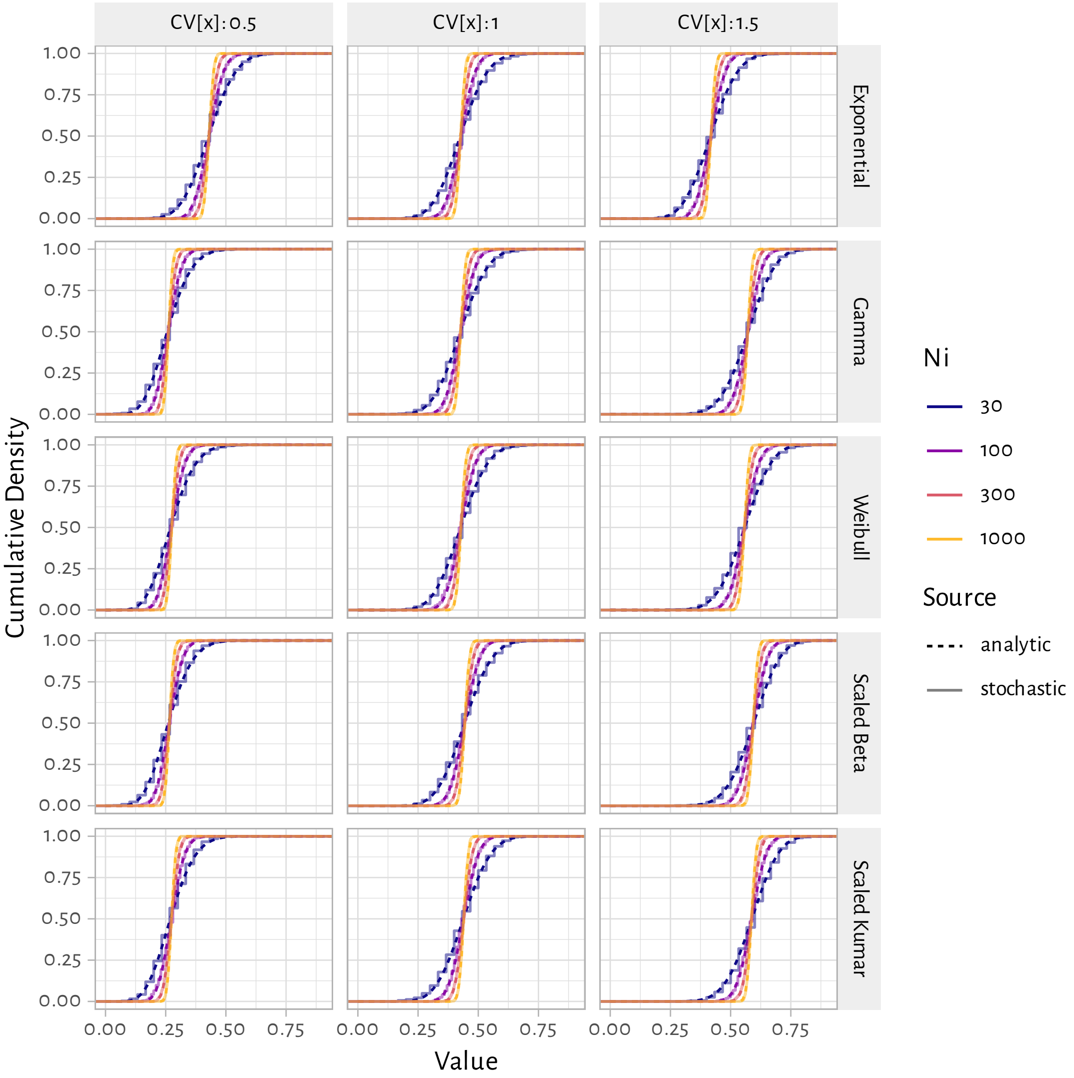


Figure A.5: Comparison of analytic likelihood functions and distribution of 10,000 stochastic estimates of $\mathbb{P}[0 < z < 1 | s]$, for different simulated survey sizes $N_i$ and latent distribution families for $f(x)$

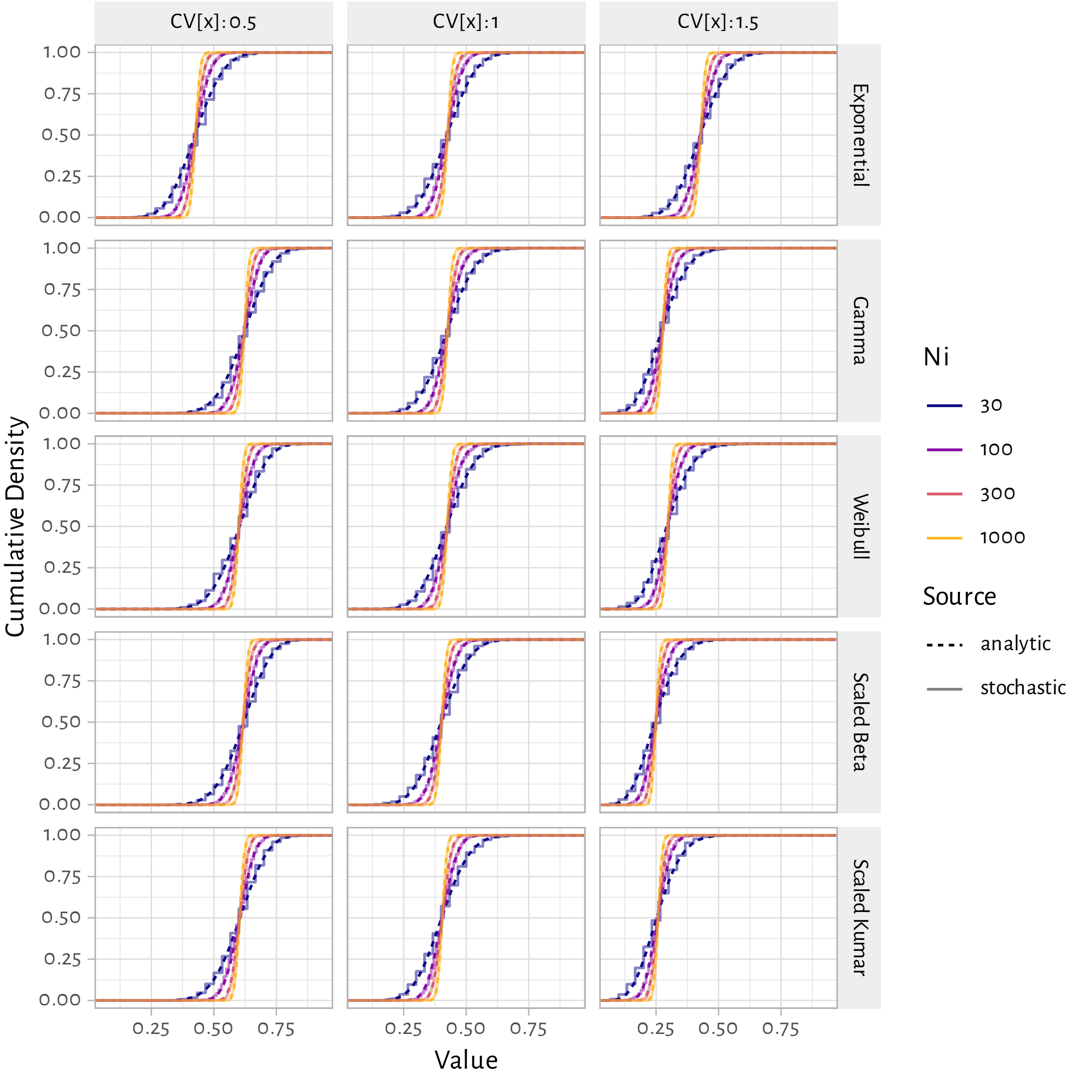


Figure A.6: Comparison of analytic likelihood functions and distribution of 10,000 stochastic estimates of $\mathbb{P}[1 < z < 5|s]$, for different simulated survey sizes $N_i$ and latent distribution families for $f(x)$

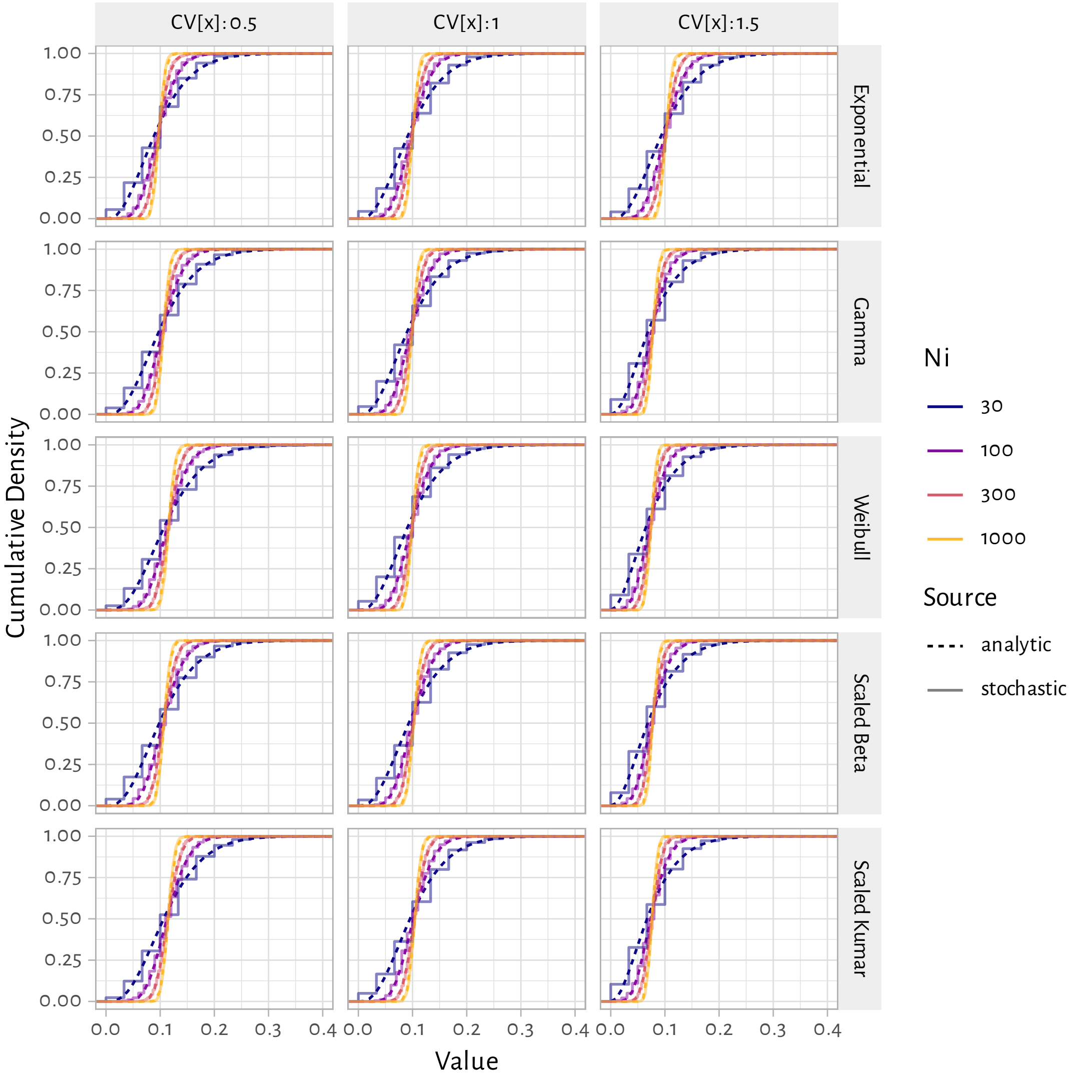


Figure A.7: Comparison of analytic likelihood functions and distribution of 10,000 stochastic estimates of $\mathbb{P}[5 < z < 9|s]$, for different simulated survey sizes $N_i$ and latent distribution families for $f(x)$

##### A.2.1 Aggregate data from a single survey

Figure A.8 illustrates the posterior distributions inferred by fitting the Bayesian model to the RDS-adjusted sex work duration data reported in [3]. The best-fitting distribution family, based on overall log-likelihood, was Gamma (purple). Figure A.9 also illustrates an example trace plot (Gamma family) showing convergence of 7 sampling chains during 500 warm-up iterations.

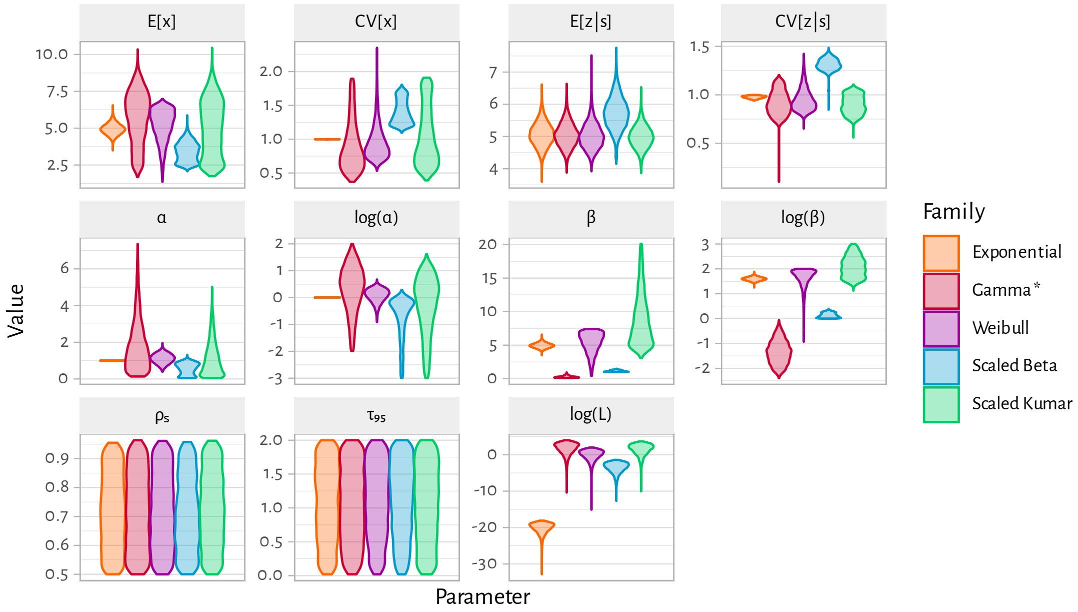


Figure A.8: Posterior distributions of model parameters and summary statistics when fitting to the RDS-adjusted Eswatini data (Table 2)

* denotes the best-fitting distribution family

##### A.2.2 Aggregate data from a global review

**Data** The manually extracted data from Table S2 in [2] are provided as supplementary file `Fazito2012.csv`. We extracted all reported duration means $\mathbb{E}[z|s]$ and interval proportions $\hat{\mathbb{P}}[a < z < b|s]$ (including medians and maximum durations), but dropped from our analyses for simplicity: standard deviations, minimum durations, and durations reported as "greater/less than X". We also dropped estimates from the following 9 sources due to the female sex worker (FSW) population definition (it's not clear whether these studies were included in the quantitative syntheses of [2]):

- Anzala (1995) Kenya: FSW in a cohort who acquired HIV
- Ward (2006) UK: FSW who retired and were followed-up
- Pineda (1995) Spain: FSW working for at least 21 months

- Strathdee (2008) Mexico: duplicate of Loza (2010) Mexico
- MOH (2003) Cambodia: not clearly FSW (e.g. karaoke workers)
- MOH (2007) Cambodia: not clearly FSW (e.g. karaoke workers)
- Gu (2009) China: FSW who inject drugs
- Silverman (2006) India: women trafficked into selling sex
- Simoes (1993) India: women incarcerated for selling sex

Before synthesizing the extracted data via the Bayesian model, we also adjusted the effective sample sizes for each study $N_i$ to reduce the influence of very large studies via:

$$N_i' = \frac{1}{\frac{1}{N_0} + \frac{1}{N_i}}, \qquad N_0 = 1000 \tag{A.4}$$

This adjustment loosely approximates a random effects approach by weighting each study more equally.

**Results** Figures A.10–A.14 illustrate the posterior distributions of model parameters, summary statistics, and cumulative/density duration distributions and means versus the data for female sex workers reviewed in [2]. Figure A.15 then summarizes estimates of mean total duration in sex work among the source population $\mathbb{E}[x]$ obtained via the Bayesian model versus those reported in [2], across all distribution families considered.

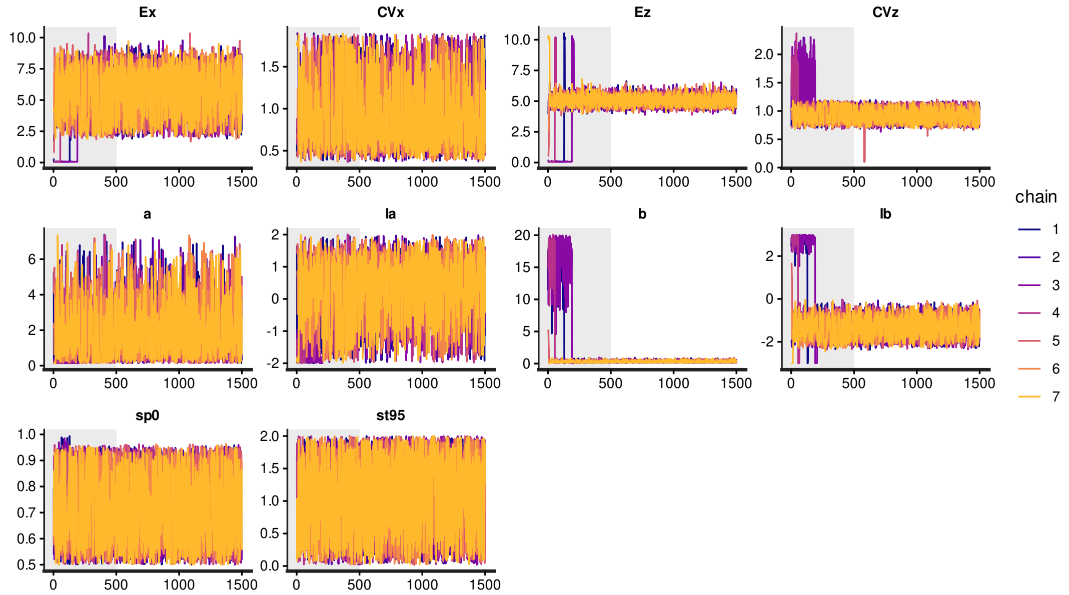


Figure A.9: Example trace plot showing convergence of 7 MCMC chains for model parameters and outcomes over 500 warm-up iterations, followed by 1000 sampling iterations

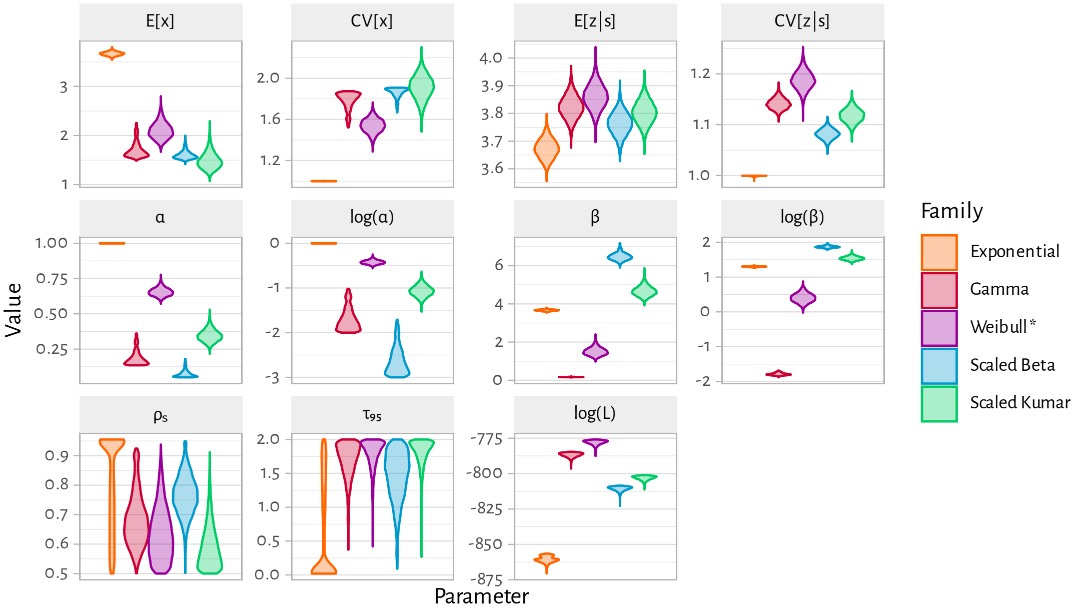


(a) Model parameter and summary statistics

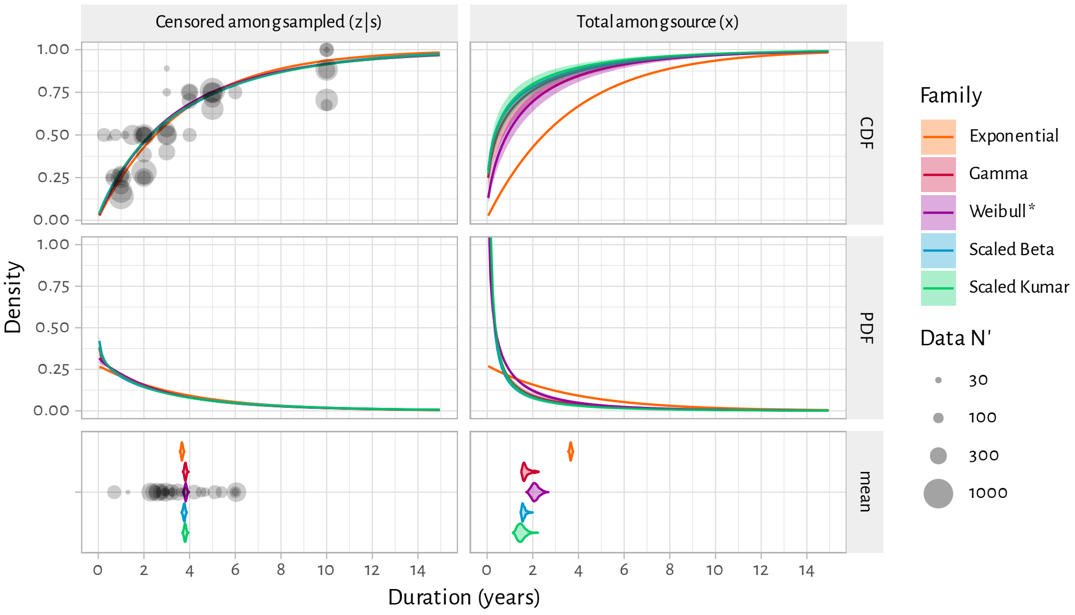


(b) Modelled cumulative (CDF) and density (PDF) distributions and means versus empirical data

Figure A.10: Bayesian model posteriors when fitting to empirical aggregate female sex worker duration data reviewed by Fazito et al. [2] for **Africa**

Coloured lines & shaded ribbons denote mean & 95% CI distribution values; gray circles denote aggregate empirical estimates; * denotes the best-fitting distribution family per region; see Table A.2 for family definitions; see Eq. (A.4) for definition of $N_i'$.

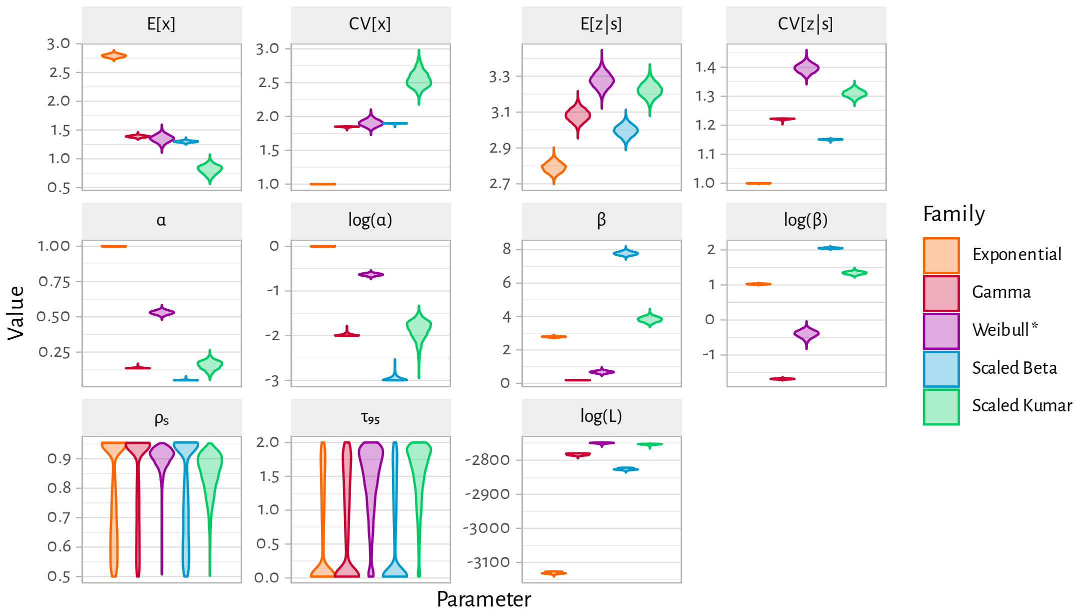


(a) Model parameter and summary statistics

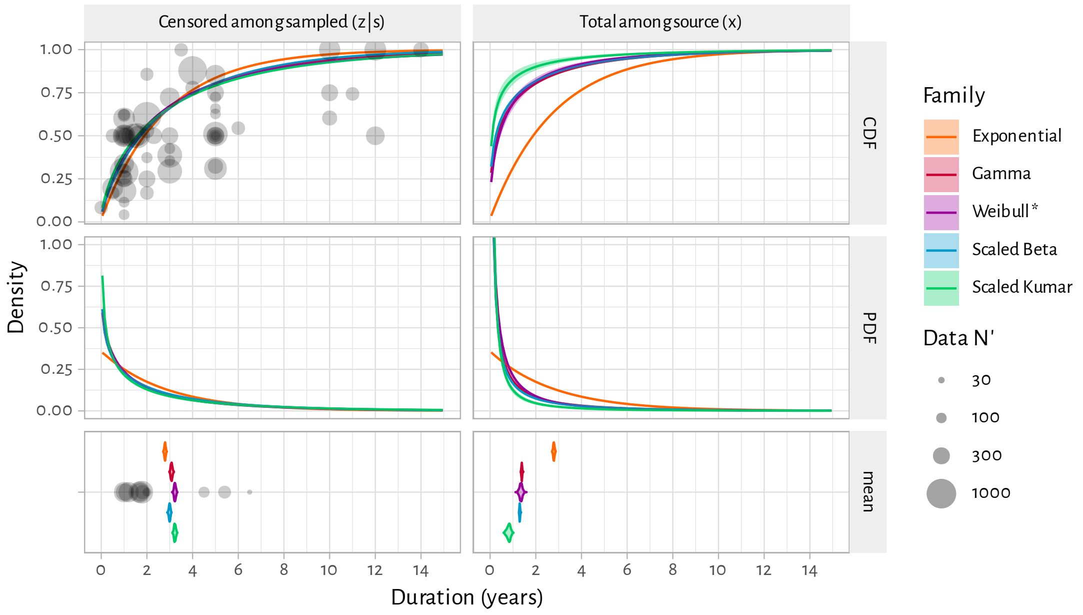


(b) Modelled cumulative (CDF) and density (PDF) distributions and means versus empirical data

Figure A.11: Bayesian model posteriors when fitting to empirical aggregate female sex worker duration data reviewed by Fazito et al. [2] for **Asia**

Coloured lines & shaded ribbons denote mean & 95% CI distribution values; gray circles denote aggregate empirical estimates; * denotes the best-fitting distribution family per region; see Table A.2 for family definitions; see Eq. (A.4) for definition of $N_i'$.

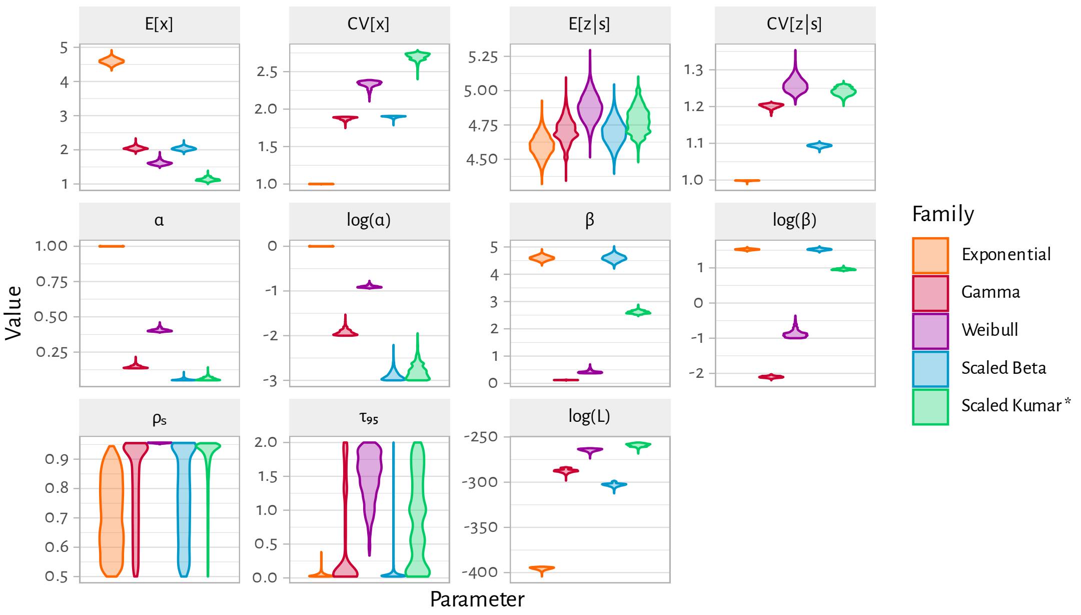


(a) Model parameter and summary statistics

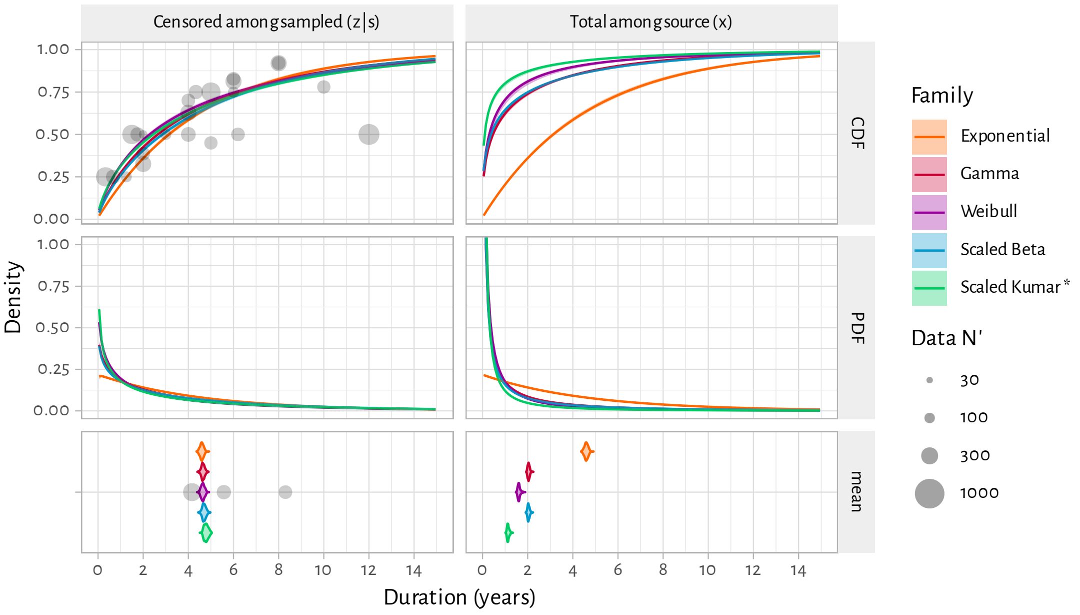


(b) Modelled cumulative (CDF) and density (PDF) distributions and means versus empirical data

Figure A.12: Bayesian model posteriors when fitting to empirical aggregate female sex worker duration data reviewed by Fazito et al. [2] for **Europe**

Coloured lines & shaded ribbons denote mean & 95% CI distribution values; gray circles denote aggregate empirical estimates; * denotes the best-fitting distribution family per region; see Table A.2 for family definitions; see Eq. (A.4) for definition of $N_i'$.

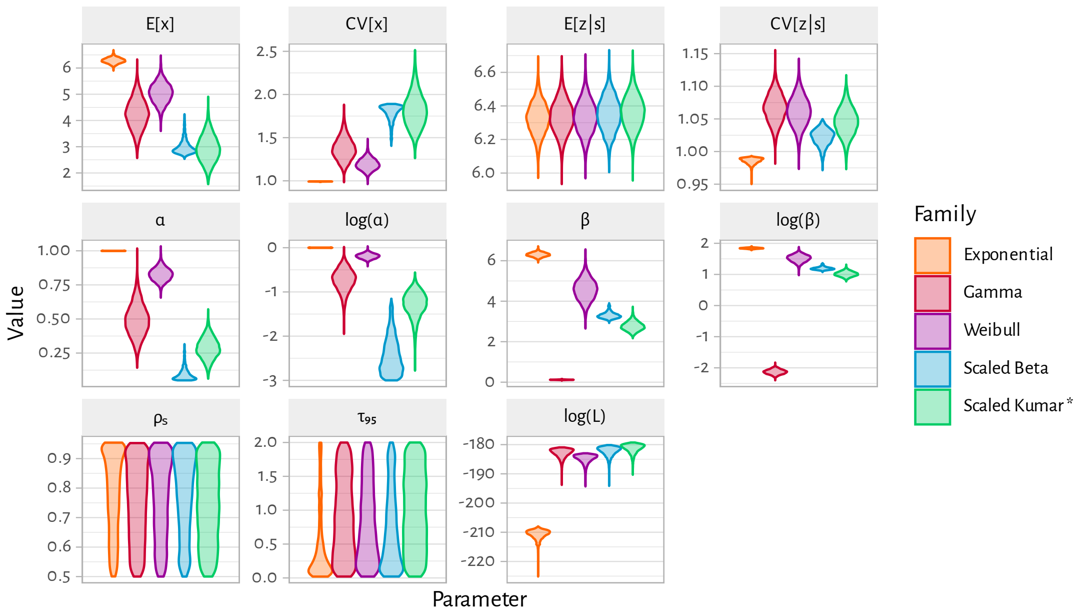


(a) Model parameter and summary statistics

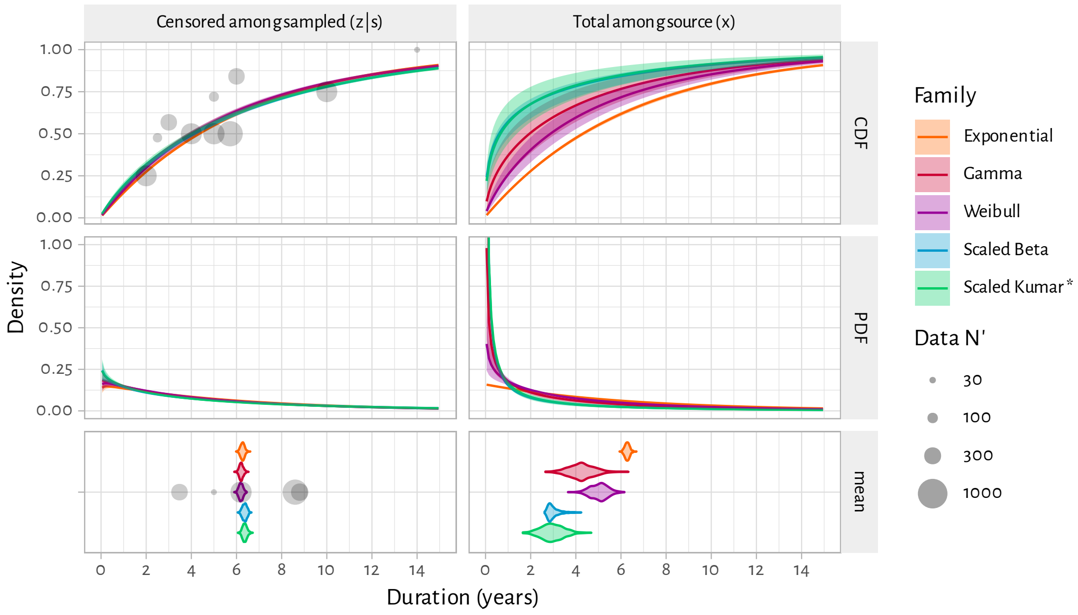


(b) Modelled cumulative (CDF) and density (PDF) distributions and means versus empirical data

Figure A.13: Bayesian model posteriors when fitting to empirical aggregate female sex worker duration data reviewed by Fazito et al. [2] for **Latin America**

Coloured lines & shaded ribbons denote mean & 95% CI distribution values; gray circles denote aggregate empirical estimates; * denotes the best-fitting distribution family per region; see Table A.2 for family definitions; see Eq. (A.4) for definition of $N_i'$.

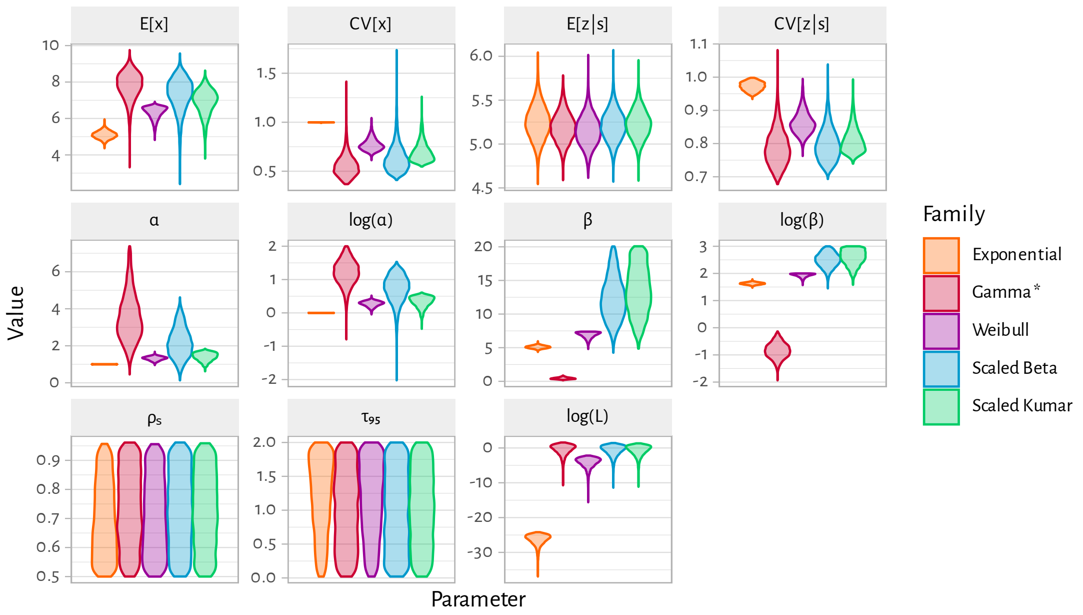


(a) Model parameter and summary statistics

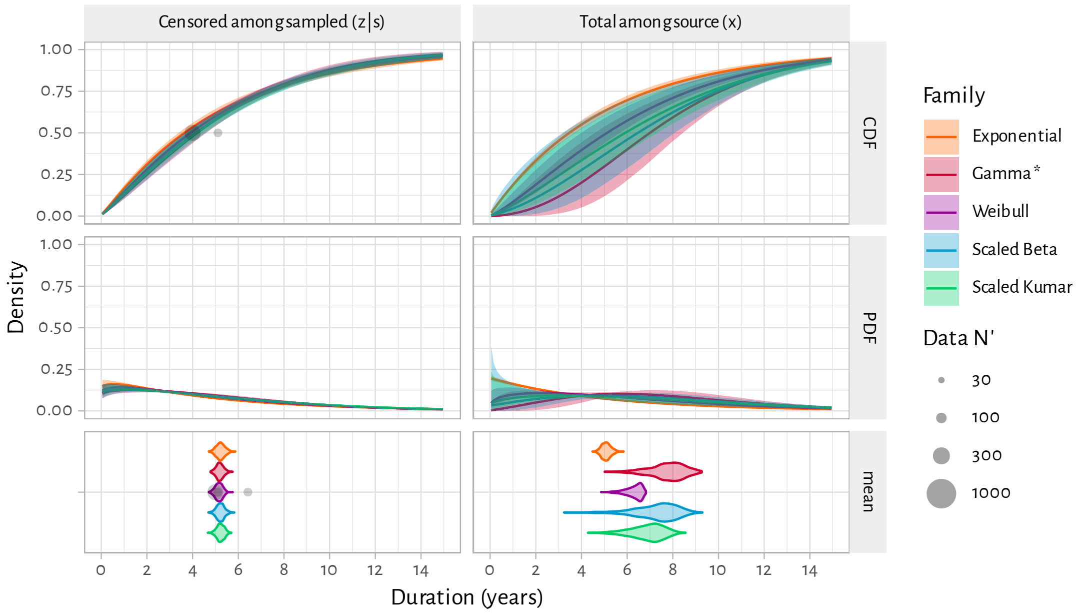


(b) Modelled cumulative (CDF) and density (PDF) distributions and means versus empirical data

Figure A.14: Bayesian model posteriors when fitting to empirical aggregate female sex worker duration data reviewed by Fazito et al. [2] for **North America**

Coloured lines & shaded ribbons denote mean & 95% CI distribution values; gray circles denote aggregate empirical estimates; * denotes the best-fitting distribution family per region; see Table A.2 for family definitions; see Eq. (A.4) for definition of $N_i$'.

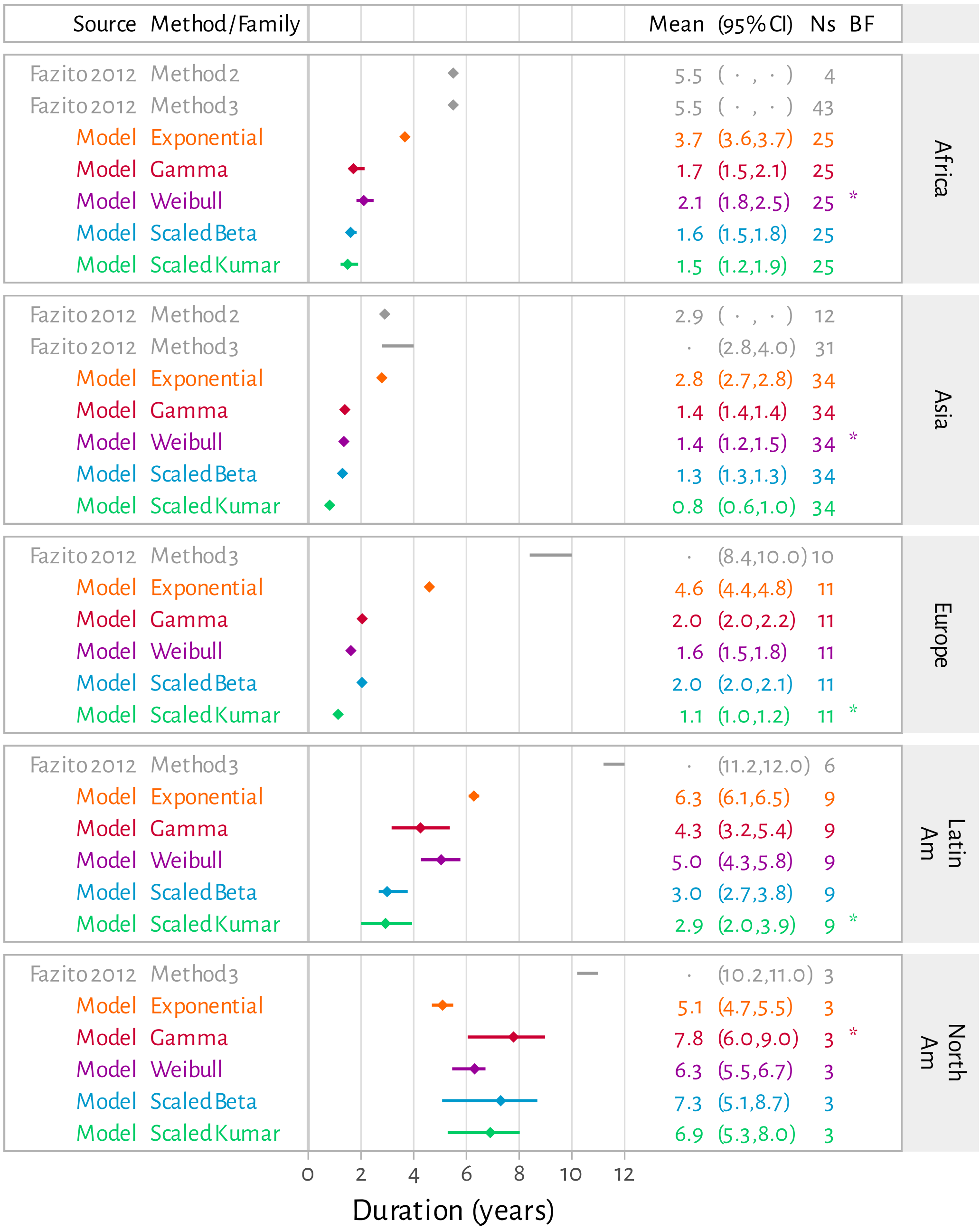


Figure A.15: Forest plot of Bayesian model-estimated mean total duration in sex work $\mathbb{E}[x]$ (all families) versus regional synthesis estimates in [2]

Synthesis methodology in [2] was unclear, and regional estimates were variably reported as points or ranges; see Table 1 for method definitions. Ns: number of unique surveys contributing to the estimate; BF: best-fitting family per region.
